# Prognostic Implications of Changes in Total Physiological Atherosclerotic Burden in Patients With Coronary Artery Disease – *A Serial Quantitative Flow Ratio Study*

**DOI:** 10.1101/2023.11.03.23298016

**Authors:** Jiapeng Chu, Deqiang Yuan, Yan Lai, Wen Ye, Lei Liu, Hao Lin, Fan Ping, Guoqi Zhu, Fei Chen, Yian Yao, Wenwen Yan, Xuebo Liu

## Abstract

The association between coronary physiological progression and clinical outcomes has not been investigated. A total of 421 patients who underwent serial coronary angiography at least 6 months apart were included. Total physiological atherosclerotic burden was characterized by sum of quantitative flow ratio in 3 epicardial vessels (3V-QFR). The relationships of the 3V-QFR and its longitudinal change (Δ3V-QFR) with major adverse cardiovascular events (MACE) were explored. 3V-QFR values derived from follow-up angiograms were slightly lower compared to baseline (2.85 [2.77, 2.90] vs. 2.86 [2.80, 2.90], *p* < 0.001). The median Δ3V-QFR value was −0.01 (−0.05, 0.02). The multivariable models demonstrated follow-up 3V-QFR or Δ3V-QFR were independently associated with MACE (both *p* < 0.05). Patients with both low follow-up 3V-QFR (≤ 2.78) and low Δ3V-QFR (≤ −0.05) presented 3 times higher risk of MACE than those without (hazard ratio: 2.953, 95% confidence interval 1.428-6.104, *p* = 0.003). Furthermore, adding patient-level 3V-QFR and Δ3V-QFR to clinical model significantly improved the predictability for MACE. In conclusion, total physiological atherosclerotic burden and its progression can provide incremental prognostic value over clinical characteristics, supporting the use of coronary physiology in the evaluation of disease progression and for the identification of vulnerable patients.

## Introduction

Coronary atherosclerosis has been thought as a systematic disease presented with a chronic and progressive process [1]. Over the last few decades, it has ignited intense clinical interest to study the natural evolution of coronary atherosclerosis, and to assess changes in plaque volume and composition in response to medical therapy using invasive or noninvasive imaging modalities, such as intravascular ultrasound (IVUS) and coronary computed tomography angiography (CTA) [2–4]. However, the clinical implication of assessing functional alterations using coronary physiological techniques has not yet been well investigated.

Fractional flow reserve (FFR) is the gold standard for determining the functional severity of coronary stenosis [5]. Although FFR is frequently used as a dichotomous value to guide coronary revascularization in clinical practice, accumulated evidence has supported the prognostic efficacy of FFR beyond a single threshold of 0.80 both before and after percutaneous coronary intervention (PCI) [6,7]. Recent studies indicate the sum of FFR values in the 3 major coronary arteries could be served as a marker of total physiological atherosclerotic burden, and could provide better risk stratification in patients with coronary artery disease (CAD) [8,9]. However, the invasive nature of FFR may pose a barrier for its routine measurements in all 3 vessels.

Quantitative flow ratio (QFR) derived from the computation of invasive coronary angiography (ICA) is an attractive alternative to wire-based FFR, allowing it possible to obtain physiological information of the entire coronary tree with no requirement for additional invasive procedures [10,11]. In this study, we aimed to explore the clinical relevance and prognostic implications of coronary functional progression by serial QFR measurements in 3 major coronary arteries.

## Methods

### Study design and population

This was a retrospective analysis of patients with suspected or diagnosed CAD who had undergone at least two ICA measurements at least 6 months apart at Tongji Hospital, Tongji University, Shanghai. The study flow is shown in **Supplementary Figure 1**. Patients with percent diameter stenosis between 20% and 90% in all 3 major epicardial arteries by visual assessment at baseline ICA and who underwent successful serial QFR measurements at both the index and follow-up procedures were enrolled. The second ICA was performed for symptomatic reasons or at the request of treating physicians for follow-up. Exclusions criteria included acute myocardial infarction (MI) within 72h at baseline, previous or planned coronary artery bypass graft surgery, significant left main disease (stenosis > 50% by visual estimation) or ostial lesions (< 3 mm from the aorta), diminutive coronary artery (proximal reference diameter < 2 mm), and patients who experienced adverse cardiac events in between the two ICA measurements or received repeated revascularization at the time of second ICA procedure. Finally, patients were excluded in cases of inadequate image quality of the angiography and severe vessel overlap or tortuosity. The study was conducted in accordance with the Declaration of Helsinki and approved by the Institutional Review Board of Tongji Hospital, Tongji University.

### Angiographic analysis

ICA and PCI were performed according to the standard of care on the basis of local practice. Quantitative coronary angiography (QCA) analyses were performed using an offline computerized quantitative coronary angiographic system (CAAS system; Pie Medical Instruments, Maastricht, The Netherlands). Minimum lumen diameter, reference diameter, lesion length, and percent diameter stenosis were routinely calculated. The anatomic severity and progression of CAD was measured via the Gensini score (GS), a widely used angiographic scoring system for quantifying coronary disease complexity [12,13]. The absolute GS change (ΔGS) was calculated by subtracting baseline GS from values obtained at follow-up. QCA and GS analyses were performed after stent implantation in vessels underwent PCI at index procedure.

### Offline QFR assessment

Offline Murray-law based QFR (μQFR), the next-generation of angiographic QFR, was performed by experienced analysts certified for the use of the QFR system software (AngioPlus Core, Pulse Medical Imaging Technology, Shanghai, China), as previously described [14]. In brief, a single angiographic projection, in which the coronary stenosis was best visualized, was transferred to the QFR system. The lumen contour of the interrogated vessel and its side branches with diameters ≥ 1 mm were automatically delineated throughout the angiographic loop. A manual correction was performed in cases of suboptimal angiographic image quality based on a standard operation procedure. A contrast flow model based on contrast flow velocity and pressure drop calculation based on fluid dynamics equations were used for QFR computations. Subsequently, the μQFR values of the interrogated vessel and its side branches were computed. QFR measurements were recorded for all 3 vessels at both index and follow-up procedures. 3V-QFR was calculated as the sum of the μQFR values measured in the 3 major coronary arteries. When a PCI was performed at the index procedure, the post-PCI QFR value was used to calculate 3V-QFR. The absolute change in 3V-QFR (Δ3V-QFR) was defined as the follow-up 3V-QFR minus the baseline 3V-QFR.

### Data collection and clinical follow-up

Baseline demographics and laboratory parameters were obtained by reviewing the hospital database. Clinical follow-up data were collected by outpatient clinic visits, hospitalization information or telephone contacts. The primary endpoint of the study was major adverse cardiac events (MACE), a composite of cardiac death, non-fatal MI, or ischemia-driven revascularization (IDR), after follow-up ICA procedure. All clinical outcomes were defined according to the Academic Research Consortium. Any death with unknown cause was regarded as cardiac in nature. The definition of MI was based on the fourth universal definition [15]. IDR was defined as a revascularization procedure with at least one of the following: (i) recurrence of angina, (ii) positive non-invasive test, and (iii) positive invasive physiologic test. Time to event was defined by the interval from the time of follow-up ICA procedure to the time of the future event.

### Statistical analysis

Continuous variables are presented as mean ± SD or median with 25% to 75% interquartile range, and compared between groups using Student’s *t* tests, Wilcoxon signed rank test or Mann–Whitney *U* test as appropriate. Categorical variables are summarized as absolute counts and percentages, and compared between groups using Pearson’s chi-square or Fisher’s exact test. Receiver operating characteristic (ROC) curve analysis was performed to assess the optimal cutoff value of 3V-QFR or Δ3V-QFR for predicting MACE. A penalized spline method was used to allow for nonlinearities in the association between continuous variables and MACE. The cumulative incidence of clinical events was presented as Kaplan-Meier estimates and compared using a log-rank test among groups classified by the cutoff values of 3V-QFR and Δ3V-QFR. Cox proportional hazards regression analysis was used to estimate hazard ratios (HR) and corresponding 95% confidence intervals (CI). Variables with a *p* value < 0.05 in univariable model were then included in multivariable Cox proportional hazard analysis. The additional prognostic values of physiological parameters over baseline clinical characteristics were assessed using ROC curve with area under curve (AUC), category-free net reclassification index (NRI) and integrated discrimination improvement (IDI). The logistic regression analysis was performed to identify the independent baseline clinical factors associated with rapid functional progression, which was defined based on the cutoff value of Δ3V-QFR (≤ −0.05). Variables with a *p* value < 0.1 in the univariable analyses or clinically relevant were entered into multivariable model. Odds ratios (OR) with corresponding 95% CI were provided. A 2-tailed *p* value < 0.05 was considered statistically significant. All statistical analyses were performed using SPSS version 26.0 (SPSS, Chicago, Illinois, USA) and R version 3.6.1 (R Foundation for Statistical Computing, Vienna, Austria).

## Results

### Patient and angiographic characteristics

Between January 2016 and December 2020, a total of 421 patients who met all clinical and imaging enrollment criteria were included in the present analysis with a median time interval of 12 (11, 14) months from baseline to secondary ICA. The baseline patient characteristics are summarized in **Table 1**. The median age was 65(59, 71) years and 304 (72.2%) were male. At baseline ICA, 249 (59.1%) patients were classified as having multivessel disease, and 385 (91.4%) patients had a history of PCI in at least 1 vessel. The baseline and follow-up angiographic and physiological characteristics are summarized in **Supplementary Table 1.** The median 3V-QFR values at follow-up were numerically lower than baseline (2.85 [2.77, 2.90] vs. 2.86 [2.80, 2.90]; *p* < 0.001 for Wilcoxon matched-pairs test). The absolute changes in QFR values on patient-level (Δ3V-QFR) and vessel-level (ΔQFR) were −0.01 (−0.05, 0.02) and 0 (−0.02, 0.01), respectively.

**Table 1.** Baseline patient characteristics (N = 421)

| Characteristics |  |
| --- | --- |
| Time interval between ICA measurements, months | 12 (11-14) |
| Age, years | 65 (59-71) |
| Male (%) | 304 (72.2) |
| Body mass index, Kg/m<sup>2</sup> | 24.9 (22.5, 26.6) |
| Current or former smoker (%) | 228 (54.2) |
| Hypertension (%) | 296 (70.3) |
| Hyperlipidemia (%) | 251 (59.6) |
| Diabetes mellitus (%) | 165 (39.2) |
| Chronic kidney disease (%) | 51 (12.1) |
| Previous myocardial infarction (%) | 89 (21.1) |
| Previous stroke (%) | 32 (7.6) |
| Clinical presentation on admission |  |
| Asymptomatic ischemia (%) | 61 (14.5) |
| Stable angina (%) | 170 (40.4) |
| Unstable angina (%) | 146 (34.7) |
| Post myocardial infarction within 1 months (%) | 44 (10.5) |
| Medication at discharge |  |
| DAPT (%) | 331 (78.6) |
| ACEI/ARB (%) | 271 (64.4) |
| B-blockers (%) | 288 (68.4) |
| Statins (%) | 413 (98.1) |
| eGFR, ml/min/1.73 m<sup>2</sup> | 83.1 (69.9, 95.5) |
| hs-CRP, mg/L | 1.74 (0.80, 3.62) |
| HbA1c, % | 6.2 (5.8, 6.9) |
| Total cholesterol, mmol/L | 4.21 (3.48, 4.91) |
| Triglycerides, mmol/L | 1.34 (1.00, 1.94) |
| HDL cholesterol, mmol/L | 1.01 (0.88, 1.19) |
| LDL cholesterol, mmol/L | 2.75 (2.11, 3.35) |
| Baseline angiographic characteristics |  |
| Multivessel disease (%) | 249 (59.1) |
| Underwent PCI (%) | 385 (91.4) |

Data are expressed as n (%), mean ± SD, or median (25th, 75th percentiles). DAPT, dual antiplatelet therapy; ACEI, angiotensin-converting enzyme inhibitor; ARB, angiotensin II receptor antagonist; eGFR, estimated glomerular filtration rate; hs-CRP, high sensitivity C-reactive protein; HbA1c, Glycosylated hemoglobin; HDL, high-density lipoprotein; LDL, low-density lipoprotein; PCI, percutaneous coronary intervention.

### Clinical outcomes and cutoff values of physiological parameters

During a median follow-up of 38 (28, 48) months after the secondary ICA, MACE was observed in 50 (11.9%) patients, including 5 cardiac death, 14 non-fatal MI and 40 IDR. Patient and angiographic characteristics stratified according to MCAE are presented in **Supplementary Table 2**. The MACE group had a significantly higher GS and 3V-QFR at both baseline and follow-up ICA, as well as lower Δ3V-QFR level than the non-MACE group. ROC curve analysis showed the cutoff value of the follow-up 3V-QFR and Δ3V-QFR for predicting subsequent MACE were 2.78 (AUC: 0.68, 95% CI: 0.60-0.76, *p* < 0.001) and −0.05 (AUC: 0.65, 95% CI: 0.57-0.74, *p* < 0.001), respectively (**Supplementary Figure 2**). When grouping the patients according to the cutoff value of the patient-level physiological indices, the log-rank test showed a significant association between a lower level of follow-up 3V-QFR or Δ3V-QFR with MACE based on the Kaplan-Meier curves (both *p* < 0.001) (**Figure 1**). The details of clinical events at 5 year from follow-up ICA according to the groups stratified by follow-up 3V-QFR or Δ3V-QFR are presented in **Table 2**. The differences in MACE between groups were mainly driven by non-fatal MI and IDR, nonsignificant difference in cardiac death was found. After adjusting for potential confounding clinical and angiographic factors, different multivariable models revealed that both follow-up 3V-QFR and Δ3V-QFR remained independently correlated with subsequent MACE. Results were unchanged when considering these two physiological indices as categorical variables to evaluate their relationships with the clinical outcomes (**Supplementary Table 3-4**).

**Figure 1.**
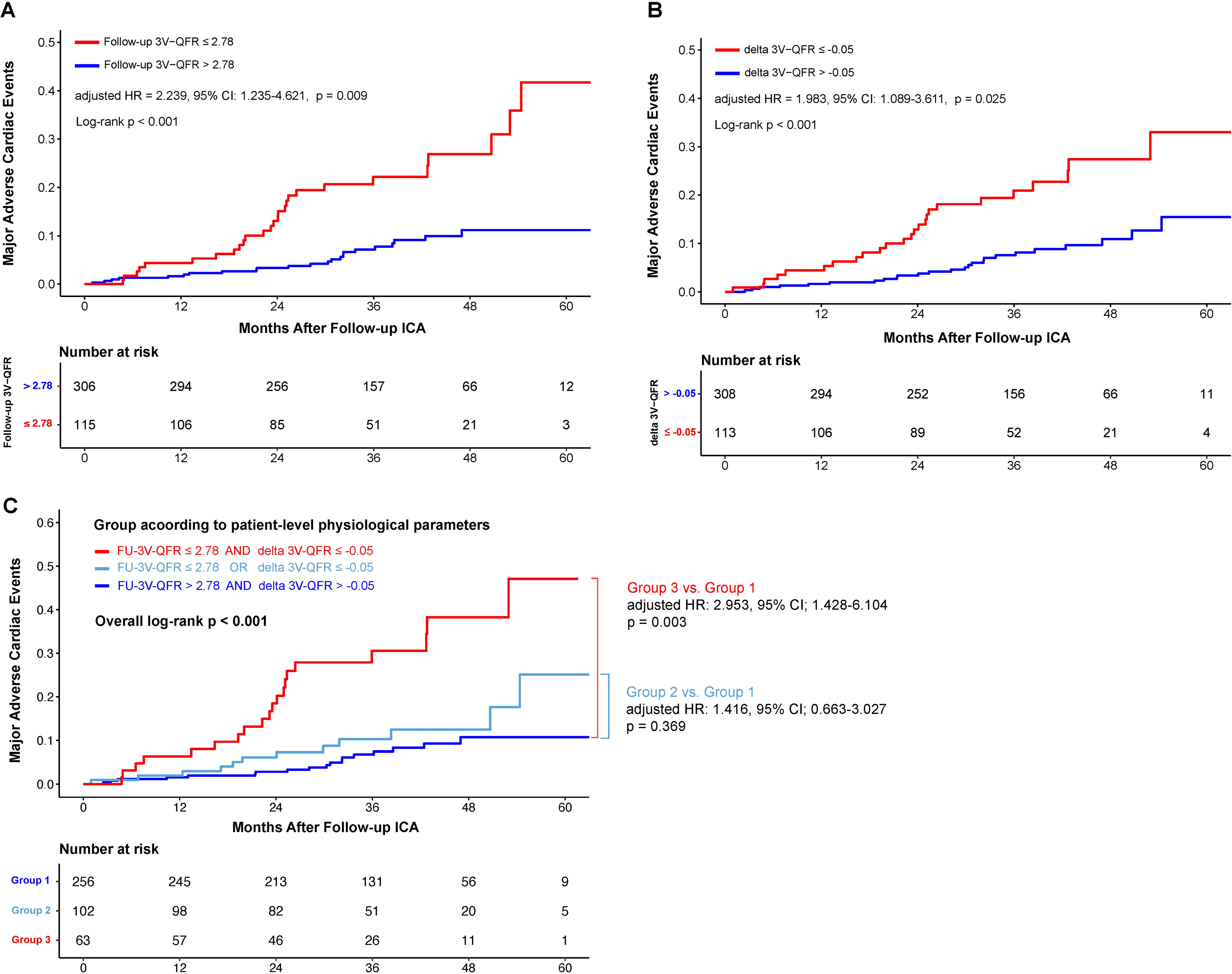
Cumulative incidence of MACE after follow-up ICA procedure according to 3V-QFR and Δ3V-QFR values. Kaplan-Meier curves of 5-year MACE are presented according to follow-up 3V-QFR (A), Δ3V-QFR (B) or both (C); MACE, major adverse cardiac events; QFR, quantitative flow ratio; HR, hazard ratio; ICA, invasive coronary angiography.

**Table 2.**
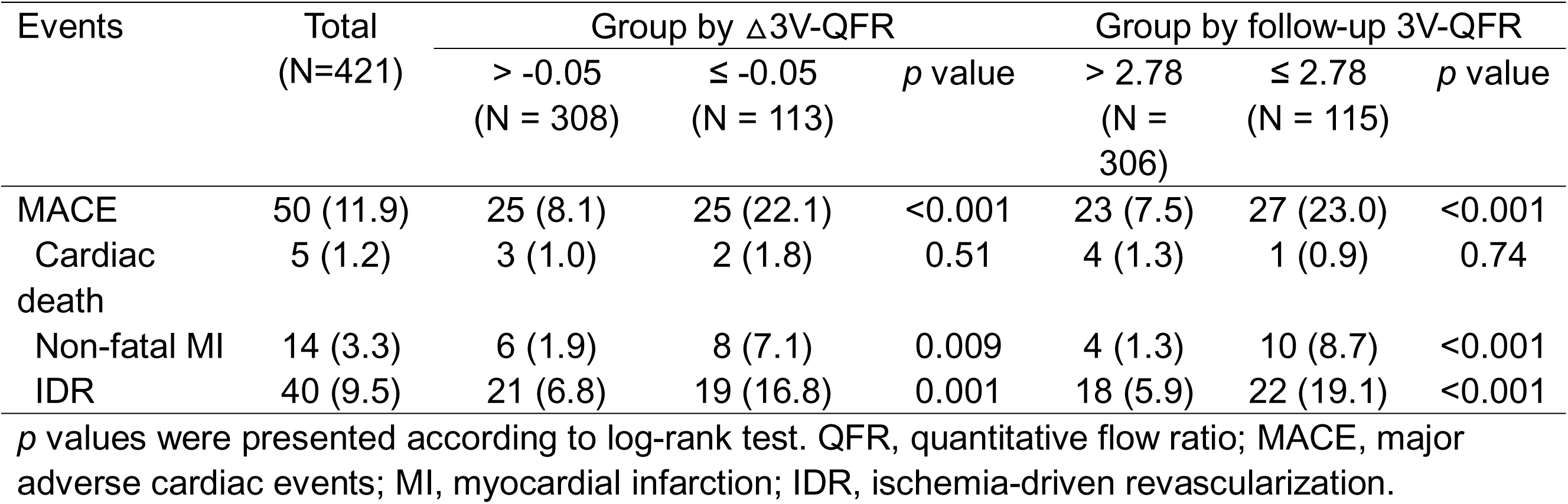
Clinical outcomes according to Δ3V-QFR or follow-up 3V-QFR.

| Events | Total<br>(N=421) | Group by Δ3V-QFR | | | Group by follow-up 3V-QFR | | |
| --- | --- | --- | --- | --- | --- | --- | --- |
| | | > -0.05<br>(N = 308) | ≤ -0.05<br>(N = 113) | *p* value | > 2.78<br>(N = 306) | ≤ 2.78<br>(N = 115) | *p* value |
| MACE | 50 (11.9) | 25 (8.1) | 25 (22.1) | <0.001 | 23 (7.5) | 27 (23.0) | <0.001 |
| Cardiac death | 5 (1.2) | 3 (1.0) | 2 (1.8) | 0.51 | 4 (1.3) | 1 (0.9) | 0.74 |
| Non-fatal MI | 14 (3.3) | 6 (1.9) | 8 (7.1) | 0.009 | 4 (1.3) | 10 (8.7) | <0.001 |
| IDR | 40 (9.5) | 21 (6.8) | 19 (16.8) | 0.001 | 18 (5.9) | 22 (19.1) | <0.001 |

*p* values were presented according to log-rank test. QFR, quantitative flow ratio; MACE, major adverse cardiac events; MI, myocardial infarction; IDR, ischemia-driven revascularization.

### Incremental prognostic value of 3V-QFR and **Δ**3V-QFR

Based on the cutoff values of follow-up 3V-QFR and Δ3V-QFR, patients were further categorized into 3 groups: Group 1: high follow-up 3V-QFR and high Δ3V-QFR; Group 2, low follow-up 3V-QFR or low Δ3V-QFR; Group 3, low follow-up 3V-QFR and low Δ3V-QFR. The cumulative incidence of 5-year MACE was 10.7%, 25.1%, and 47.1% for Group 1 to Group 3, respectively (overall log-rank *p* < 0.001). In adjusted analysis, patients in the Group 3 had a 3-fold increased risk of MACE compared with Group 1 (HR: 2.953, 95% CI, 1.428 to 6.104, *p* = 0.003) (**Figure 1C**). Compared with the model consisted of clinical and angiographic risk factors, adding groups categorized by patient-level physiological parameters (follow-up 3V-QFR ≤ 2.78 and Δ3V-QFR ≤ −0.05) significantly improved the discriminatory ability (AUC: 0.77 vs. 0.72, difference in AUC: 0.05, *p* = 0.03) and reclassification ability (NRI: 0.57, *p* < 0.001; IDI: 0.03, *p* = 0.04) (**Figure 2**).

**Figure 2.**
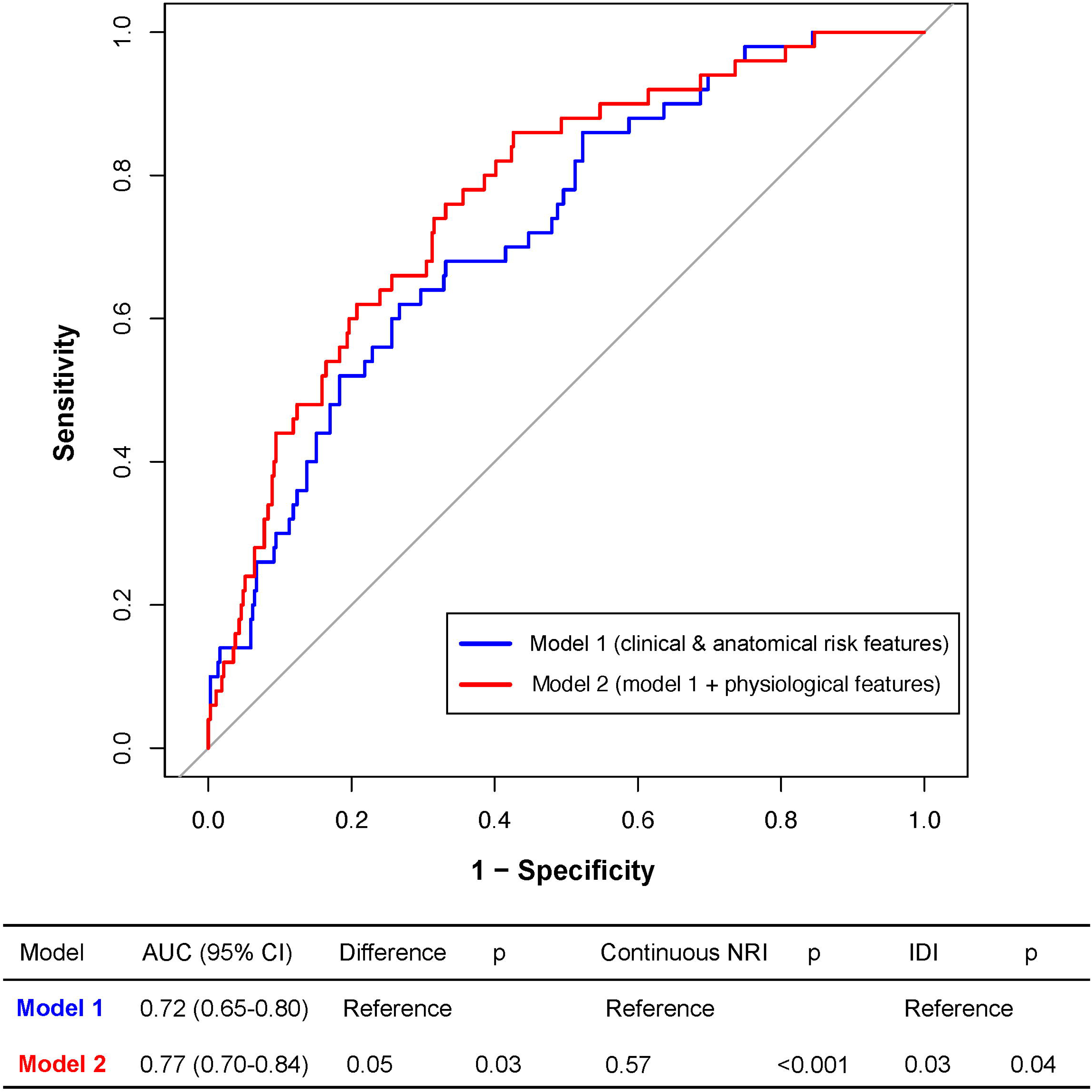
Discriminatory and reclassification ability of predictive models for future MACE. Compared with model 1 included clinical and anatomic risk factors only (age, body mass index, diabetes mellitus, history of smoking, hypertension, chronic kidney disease, acute coronary syndrome, multivessel disease at baseline as well as follow-up GS and ΔGS), predictive model 2 with patient-level physiological parameters (follow-up 3V-QFR and Δ3V-QFR) showed a significantly higher discriminatory and reclassification abilities in identifying patients with subsequent MACE. QFR, quantitative flow ratio; AUC, area under curve; CI: confidence interval; NRI, net reclassification index; IDI, integrated discrimination improvement; GS, Gensini score.

### Clinical factors associated with coronary functional progression

The patient characteristics stratified by the cutoff value of Δ3V-QFR are summarized in **Supplementary Table 5**. The patients with lower Δ3V-QFR (≤ −0.05) had significantly higher prevalence of prior history of MI, higher proportion of renin angiotensin aldosterone system inhibitors use at discharge, and higher incidence of multivessel disease than those with Δ3V-QFR > −0.05. The high sensitivity C-reactive protein, total cholesterol and low-density lipoprotein (LDL) cholesterol levels were also higher in patients with Δ3V-QFR ≤ −0.05. Multivariable logistic regression analysis showed that previous MI and baseline LDL cholesterol were independent clinical predictors for low Δ3V-QFR (**Table 3**).

**Table 3.**
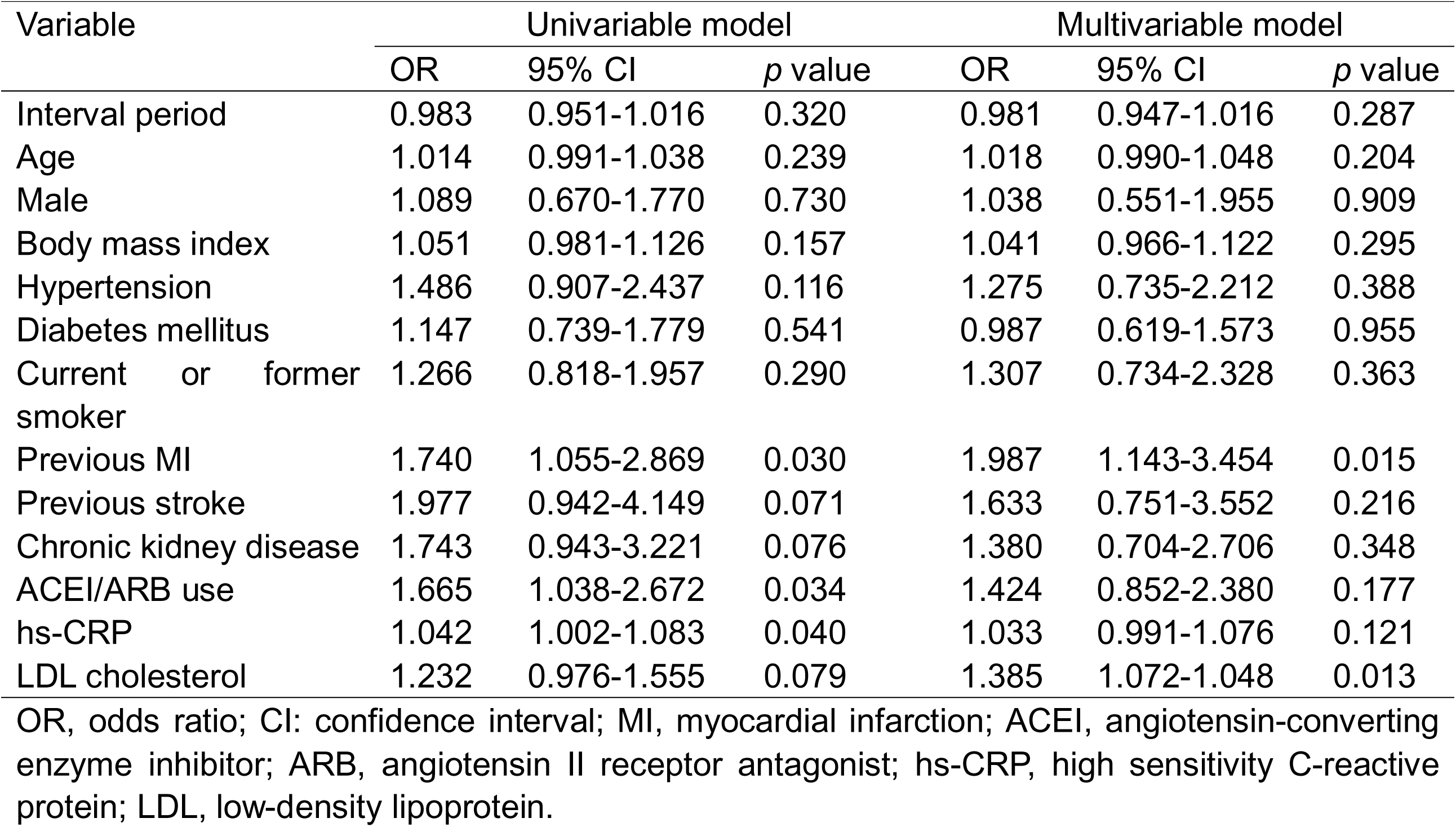
Multivariable logistic regression analysis for baseline clinical factors associated with low.

| Variable | Univariable model |  |  | Multivariable model |  |  |
| --- | --- | --- | --- | --- | --- | --- |
|  | OR | 95% CI | *p* value | OR | 95% CI | *p* value |
| Interval period | 0.983 | 0.951-1.016 | 0.320 | 0.981 | 0.947-1.016 | 0.287 |
| Age | 1.014 | 0.991-1.038 | 0.239 | 1.018 | 0.990-1.048 | 0.204 |
| Male | 1.089 | 0.670-1.770 | 0.730 | 1.038 | 0.551-1.955 | 0.909 |
| Body mass index | 1.051 | 0.981-1.126 | 0.157 | 1.041 | 0.966-1.122 | 0.295 |
| Hypertension | 1.486 | 0.907-2.437 | 0.116 | 1.275 | 0.735-2.212 | 0.388 |
| Diabetes mellitus | 1.147 | 0.739-1.779 | 0.541 | 0.987 | 0.619-1.573 | 0.955 |
| Current or former smoker | 1.266 | 0.818-1.957 | 0.290 | 1.307 | 0.734-2.328 | 0.363 |
| Previous MI | 1.740 | 1.055-2.869 | 0.030 | 1.987 | 1.143-3.454 | 0.015 |
| Previous stroke | 1.977 | 0.942-4.149 | 0.071 | 1.633 | 0.751-3.552 | 0.216 |
| Chronic kidney disease | 1.743 | 0.943-3.221 | 0.076 | 1.380 | 0.704-2.706 | 0.348 |
| ACEI/ARB use | 1.665 | 1.038-2.672 | 0.034 | 1.424 | 0.852-2.380 | 0.177 |
| hs-CRP | 1.042 | 1.002-1.083 | 0.040 | 1.033 | 0.991-1.076 | 0.121 |
| LDL cholesterol | 1.232 | 0.976-1.555 | 0.079 | 1.385 | 1.072-1.048 | 0.013 |

OR, odds ratio; CI: confidence interval; MI, myocardial infarction; ACEI, angiotensin-converting enzyme inhibitor; ARB, angiotensin II receptor antagonist; hs-CRP, high sensitivity C-reactive protein; LDL, low-density lipoprotein.

## Discussion

To the best of our knowledge, this is the first study to evaluate the clinical relevance of total physiological burden progression by serial QFR measurements in 3 coronary arteries. The major findings of this study include: (1) The total physiological burden showed a slow progression rate, as assessed by 3V-QFR at a median value of 0.01 decrease during 1-year interval; (2) Patient-level 3V-QFR and Δ3V-QFR were independently associated with subsequent risk of MACE. Integration of these physiological features into clinical model significantly improved discrimination and reclassification abilities for MACE; and (3) Previous MI and baseline LDL cholesterol were independently associated with rapid coronary functional progression defined by Δ3V-QFR ≤ −0.05.

A variety of coronary imaging modalities have been developed in recent years to precisely assess atherosclerotic plaque. As traditional ICA only provides information regarding the vessel lumen, novel imaging strategies such as coronary CTA and IVUS have emerged as the main tools for assessing plaque volume and composition, and monitoring its dynamic change in response to medical treatments [16]. However, the clinical utility of coronary physiology in assessing atherosclerosis progression remains limited. In daily practice, physiological measurements obtained during invasive procedures have been used as a dichotomous index to identify flow-limiting stenosis (i.e., FFR ≤ 0.80) requiring coronary revascularization [5,17]. Indeed, these coronary physiological parameters, as a continuous value, allow the assessment of functional severity or ischemic burden of coronary stenosis [6]. Building on this concept, Lee et al. [8] proposed the sum of FFR values of 3 major coronary arteries as a surrogate marker of total physiological atherosclerotic burden and demonstrated its influence on clinical outcomes among 1136 CAD patients. However, routine measurement of hemodynamic status in all 3 vessels using invasive FFR is not feasible due to a series of concerns, including increased procedural time and cost, the need for pressure wire advancement and pharmacologic hyperemia induction. From a practical standpoint, angiography-derived QFR, which can be easily obtained with no requirement for additional invasive procedures, allows serial evaluation of ischemic burden from the entire coronary tree [11,14].

In the present study, the novel generation of the QFR system was used for the serial functional assessment, and the analyzability of 3V-QFR was acceptable. Based on our patient cohort, total physiological atherosclerotic burden assessed by 3V-QFR at baseline or follow-up was significantly higher in those experiencing MACE, which is consistent with prior observational 3-vessel FFR or QFR studies [8,9,18]. Of importance, we also found patient-level functional progression defined by absolute change in 3V-QFR ≤ −0.05, in addition to physiological atherosclerotic burden, was also independently associated with mid-term clinical outcomes in CAD patients. Accumulating serial coronary imaging studies have reported the prognostic value of IVUS or CCTA-derived assessments of patient-level disease progression for predicting future adverse events [19,20]. In line with these findings, our results provided further evidence from a physiological aspect and we also revealed the additional prognostic value of total physiological atherosclerotic burden and progression for risk stratification, supporting the use of angiography-based QFR as a useful imaging approach for the identification of “vulnerable” patients.

The longitudinal change in coronary physiology was firstly explored by Xaplanteris et al. [21] using serial FFR measurements, and the results showed that the functional progression rate is slow, as evaluated by vessel-level FFR with a median decrease of 0.007 per year. We found in our cohort that the total physiological atherosclerosis burden quantified by patient-level 3V-QFR also declined at a slow rate (median Δ3V-QFR: 0.01 during 1-year time interval). Indeed, the rate of functional progression in our study seems even slower than the aforementioned longitudinal FFR study on a vessel basis. This difference may partly due to lower angiographic lesion severity and lower risk among patients included in our study. In addition, it has been well-acknowledged that disease progression is a modifiable step between early atherosclerosis and acute coronary events. Many studies have reported that multiple pharmacological treatments, i.e., intensive lipid-lowing therapy with statins, can halt disease progression of coronary atherosclerosis [16,22]. Therefore, several studies have investigated the clinical factors associated with coronary physiological progression and the effect of conventional lipid-lowing therapies on hemodynamic status. A recent serial coronary CTA study by Yu et al. [23], on the basis of 153 low-risk patients with mild to intermediate coronary stenosis, suggested that rosuvastatin treatment could improve hemodynamic status for non-calcified plaques assessed by lesion-specific CT-derived FFR. Another 2 recently published serial QFR studies showed that a close relationship exists between the on-treatment LDL cholesterol level and coronary functional progression as evaluated by per-vessel QFR in patients with residual stenosis after PCI [24,25]. These results support the benefits of intensive lipid-lowing therapies on atherosclerosis regression from a physiological aspect. In the current study enrolled patients with established CAD and received secondary prevention, we found that baseline LDL cholesterol level and the presence of MI were independently associated with accelerated progression of total physiological atherosclerosis burden. This further suggest the importance of cardiovascular risk factor control in individuals despite already treated with interventional and statin therapies, and implied that serial evaluation of coronary physiological changes using angiographic QFR could be used as a surrogate marker for monitoring therapeutic responses in patients with CAD, especially for those high-risk patients with MI.

Several limitations should be taken into account. First, this was a single-center, retrospective observational study, the relatively small population enrolled due to rigorous inclusion and exclusion criteria, which might introduce a selection bias. Conclusions drawn from the findings cannot be extrapolated to the excluded patients. Second, the change in patient-level physiological burden was evaluated by angiographic QFR instead of pressure wire-based FFR. Nevertheless, QFR bears the potential for wider application in this clinical setting due to its less-invasive nature. Third, only patients who did not experienced adverse events during the ICA measurements were included in the analysis, which might have led to an underestimation of the progression rate of coronary physiology. In addition, the optimal threshold values for identifying significant coronary functional progression may vary by study population with a different clinical background. Future prospective studies are needed to further address this issue. Fourth, a large proportion (91.4%) of the patients underwent PCI in at least 1 coronary artery. Therefore, our data do not represent the natural history of the physiological progression of atherosclerosis disease. Finally, invasive intravascular imaging modalities such as IVUS or optical coherence tomography which may provide more information about plaque burden or vulnerable characteristics were not performed in this study.

## Conclusion

High total physiological atherosclerotic burden assessed by 3V-QFR and its progression were strongly associated with subsequent adverse cardiovascular events in patients with CAD, supporting the clinical value of angiography-based physiology in the identification of “vulnerable” patients who may derive benefits from intensive anti-atherosclerosis treatments.

## Data Availability

All data produced in the present study are available upon reasonable request to the authors

