## Supplementary Materials for "Prognostic Implications of Changes in Total Physiological Atherosclerotic Burden in Patients With Coronary Artery Disease – *A Serial Quantitative Flow Ratio Study*"


Jiapeng Chu, MD [1], Deqiang Yuan, MD [1], Yan Lai, MD [1], Wen Ye, MD [1], Lei Liu, MD [1], Hao Lin, MD [1], Fan Ping, MD [1], Guoqi Zhu, MD [1], Fei Chen, PhD [1], Yian Yao, MD [1], Wenwen Yan, PhD [1,#], Xuebo Liu, PhD [1,#]

**Affiliations:**

[1] Department of Cardiology, Tongji Hospital, School of Medicine, Tongji University, Shanghai, China.

**[#]Corresponding authors:**

Xuebo Liu, PhD, No.389, Xincun Rd, Putuo District, Shanghai 200065, China.; Tel: +86 21-66111682, Fax: +86 21-66111682\

Wenwen Yan, PhD, No.389, Xincun Rd, Putuo District, Shanghai 200065, China..

**Supplementary Table 1.** Baseline and follow-up angiographic and physiological characteristics

| | Baseline | Follow-up | *p* value |
|---|---|---|---|
| Patient-level analysis | | | |
| Gensini score | 21.0 (15.0, 29.0) | 22.5 (15.5, 30.5) | <0.001 |
| 3V-QFR | 2.86 (2.80, 2.90) | 2.85 (2.77, 2.90) | <0.001 |
| Any vessel QFR ≤ 0.80 (%) | 28 (6.7) | 37 (8.8) | 0.245 |
| Vessel-level analysis | | | |
| Minimal lumen diameter, mm | 2.2 (1.8, 2.5) | 2.0 (1.7, 2.4) | <0.001 |
| Reference diameter, mm | 3.0 (2.5, 3.5) | 2.9 (2.5, 3.4) | <0.001 |
| Diameter stenosis, % | 25 (21, 32) | 27 (21, 34) | <0.001 |
| Lesion length, mm (%) | 11.6 (7.4, 18.1) | 11.9 (7.6, 18.5) | 0.647 |
| Vessel QFR | 0.96 (0.93, 0.98) | 0.96 (0.92, 0.98) | <0.001 |
| QFR ≤ 0.80 (%) | 29 (2.3) | 40 (3.2) | 0.179 |

Data are expressed as n (%), mean ± SD, or median (25th, 75th percentiles). Continuous variables were compared between baseline and follow-up using Wilcoxon signed rank test and corresponding p-values are presented. QFR, quantitative flow ratio.

**Supplementary Table 2**. Patient characteristics stratified according to the presence of MACE

| | No MACE (N=371) | MACE (N=50) | *p* value |
|---|---|---|---|
| Time interval between ICA measurements, months | 12.1 (10.7, 14.2) | 11.9 (9.0, 15.8) | 0.267 |
| Age, years | 64 (59, 71) | 67 (60, 75) | 0.105 |
| Male (%) | 264 (71.2) | 40 (80.0) | 0.192 |
| Body mass index, $Kg/m^2$ | 24.8 (22.5, 26.5) | 25.1 (23.7, 26.8) | 0.167 |
| Current or former smoker (%) | 195 (52.6) | 33 (66.0) | 0.073 |
| Hypertension (%) | 258 (69.5) | 38 (76.0) | 0.348 |
| Hyperlipidemia (%) | 221 (59.6) | 30 (60.0) | 0.953 |
| Diabetes mellitus (%) | 137 (36.9) | 28 (56.0) | 0.010 |
| Chronic kidney disease (%) | 42 (11.3) | 9 (18.0) | 0.174 |
| Previous myocardial infarction (%) | 78 (21.0) | 11 (22.0) | 0.874 |
| Previous stroke (%) | 28 (7.5) | 4 (8.0) | 0.782 |
| Clinical presentation on admission | | | 0.155 |
| Asymptomatic ischemia (%) | 58 (15.6) | 3 (6.0) | |
| Stable angina (%) | 150 (40.4) | 20 (40.0) | |
| Unstable angina (%) | 123 (33.2) | 23 (46.0) | |
| Post myocardial infarction within 1 months (%) | 40 (10.8) | 4 (8.0) | |
| Baseline angiographic characteristics | | | |
| Multivessel disease (%) | 210 (56.6) | 39 (78.0) | 0.004 |
| Underwent PCI (%) | 338 (91.1) | 47 (94.0) | 0.786 |
| Medication at discharge | | | |
| Dual antiplatelet therapy (%) | 293 (79.0) | 38 (76.0) | 0.630 |
| ACEI/ARB (%) | 239 (64.4) | 32 (64.0) | 0.954 |
| B-blockers (%) | 253 (68.2) | 35 (70.0) | 0.797 |
| Statins (%) | 365 (98.4) | 48 (96.0) | 0.244 |
| Estimated glomerular filtration rate, ml/min/1.73 $m^2$ | 84.3 (70.5, 95.5) | 78.3 (66.6, 96.5) | 0.320 |
| high sensitivity C-reactive protein, mg/L | 1.69 (0.80, 3.38) | 2.09 (0.86, 4.24) | 0.400 |
| Glycosylated hemoglobin, % | 6.1 (5.8, 6.8) | 6.5 (6.0, 7.8) | 0.016 |
| Total cholesterol, mmol/L | 4.24 (3.48, 4.97) | 4.02 (3.50, 4.75) | 0.410 |
| Triglycerides, mmol/L | 1.37 (1.00, 1.92) | 1.24 (0.93, 2.11) | 0.915 |
| High-density lipoprotein cholesterol, mmol/L | 1.01 (0.88, 1.19) | 1.01 (0.86,1.17) | 0.550 |
| Low-density lipoprotein cholesterol, mmol/L | 2.79 (2.11, 3.38) | 2.63 (2.12, 3.18) | 0.453 |
| Baseline GS | 20.5 (14.5, 28.5) | 25.0 (20.0, 30.0) | 0.032 |
| Follow-up GS | 22.0 (15.5, 29.5) | 27.0 (22.0, 34.0) | 0.005 |
| △GS | 0.5 (0, 2.0) | 1.5 (0, 3.5) | 0.061 |
| Baseline 3V-QFR | 2.87 (2.80, 2.91) | 2.84 (2.78, 2.88) | 0.016 |
| Follow-up 3V-QFR | 2.85 (2.79, 2.90) | 2.77 (2.72, 2.88) | <0.001 |
| △3V-QFR | -0.01 (-0.04, 0.02) | -0.05 (-0.09, 0) | <0.001 |

Data are expressed as n (%), mean ± SD, or median (25th, 75th percentiles). MACE, major adverse cardiac event; PCI, percutaneous coronary intervention; ACEI, angiotensin-converting enzyme inhibitor; ARB, angiotensin II receptor antagonist; GS, Gensini score; QFR, quantitative flow ratio.

**Supplementary Table 3**. Multivariable cox regression analyses for follow-up 3V-QFR predicting MACE

| | Continuous (per 0.01 increase in follow-up 3V-QFR) | | | Category (Follow-up 3V-QFR ≤ 2.78 vs. > -2.78) | | |
|---|---|---|---|---|---|---|
| | HR | 95% CI | *p* value | HR | 95% CI | *p* value |
| Unadjusted | 0.952 | 0.933-0.971 | <0.001 | 3.490 | 2.000-6.088 | <0.001 |
| Model 1 | 0.961 | 0.939-0.984 | 0.001 | 2.578 | 1.408-4.722 | 0.002 |
| Model 2 | 0.968 | 0.941-0.997 | 0.031 | 2.389 | 1.235-4.621 | 0.010 |

Values are presented as HRs (with 95% CIs) derived via Cox proportional hazard regression analysis. Model 1: adjusted for age, body mass index, diabetes mellitus, history of smoking and multivessel disease at baseline. Model 2: Model 1 + adjusted for Gensini score calculated based on baseline and follow-up coronary angiograms. QFR, quantitative flow ratio; HR, hazard ratio; CI: confidence interval.

**Supplementary Table 4.** Multivariable cox regression analyses for △3V-QFR predicting MACE

| | Continuous (per 0.01 increase in △3V-QFR) | | | Category (△3V-QFR ≤ -0.05 vs. > -0.05) | | |
|---|---|---|---|---|---|---|
| | HR | 95% CI | *p* value | HR | 95% CI | *p* value |
| Unadjusted | 0.914 | 0.883-0.946 | <0.001 | 2.928 | 1.681-5.098 | <0.001 |
| Model 1 | 0.924 | 0.890-0.960 | <0.001 | 2.545 | 1.452-4.458 | 0.001 |
| Model 2 | 0.949 | 0.902-0.998 | 0.043 | 1.983 | 1.089-3.611 | 0.025 |

Values are presented as HRs (with 95% CIs) derived via Cox proportional hazard regression analysis. Model 1: adjusted for age, body mass index, diabetes mellitus, history of smoking and multivessel disease at baseline. Model 2: Model 1 + adjusted for Gensini score calculated from baseline and follow-up angiograms. QFR, quantitative flow ratio; HR, hazard ratio; CI: confidence interval.

**Supplementary Table 5.** Characteristics of patients stratified by the cutoff value of △3V-QFR

| | △3V-QFR > -0.05 (N=308) | △3V-QFR ≤ -0.05 (N=113) | p-value |
|---|---|---|---|
| Time interval between ICA measurements, months | 12.1 (10.5, 14.3) | 12.1(10.9, 14.3) | 0.952 |
| Age, years | 65 (59, 71) | 65 (60, 71) | 0.375 |
| Male (%) | 221 (71.8) | 83 (73.5) | 0.730 |
| Body mass index, Kg/m$^2$ | 24.7 (22.2, 26.5) | 25.2 (23.2, 26.8) | 0.069 |
| Current or former smoker (%) | 162 (52.6) | 66 (58.4) | 0.289 |
| Hypertension (%) | 210 (68.2) | 86 (76.1) | 0.115 |
| Hyperlipidemia (%) | 180 (58.4) | 71 (62.8) | 0.416 |
| Diabetes mellitus (%) | 118 (38.3) | 47 (41.6) | 0.541 |
| Chronic kidney disease (%) | 32 (10.4) | 19 (16.8) | 0.073 |
| Previous myocardial infarction (%) | 57 (18.5) | 32 (28.3) | 0.029 |
| Previous stroke (%) | 19 (6.2) | 13 (11.5) | 0.067 |
| Clinical presentation on admission | | | 0.854 |
| Asymptomatic ischemia (%) | 45 (14.6) | 16 (14.2) | |
| Stable angina (%) | 121 (39.3) | 49 (43.4) | |
| Unstable angina (%) | 108 (35.1) | 38 (33.6) | |
| Post myocardial infarction within 1 months (%) | 34 (11.0) | 10 (8.8) | |
| Baseline angiographic characteristics | | | |
| Multivessel disease (%) | 172 (55.8) | 77 (68.1) | 0.023 |
| Underwent PCI (%) | 281 (91.2) | 104 (92.0) | 0.794 |
| Medication at discharge | | | |
| Dual antiplatelet therapy (%) | 242 (78.6) | 89 (78.8) | 0.966 |
| ACEI/ARB (%) | 189 (61.4) | 82 (72.6) | 0.033 |
| B-blockers (%) | 206 (66.9) | 82 (72.6) | 0.266 |
| Statins (%) | 301 (97.7) | 112 (99.1) | 0.688 |
| Estimated glomerular filtration, ml/min/1.73 m$^2$ | 84.5 (71.6, 95.5) | 80.6 (66.2, 95.2) | 0.102 |
| high sensitivity C-reactive protein, mg/L | 1.61 (0.74, 3.05) | 2.04(1.05, 5.29) | 0.026 |
| Glycosylated hemoglobin, % | 6.1 (5.8, 6.8) | 6.3(5.9, 7.1) | 0.131 |
| Total cholesterol, mmol/L | 4.15 (3.40, 4.86) | 4.42(3.66, 5.07) | 0.048 |
| Triglycerides, mmol/L | 1.32 (0.98, 1.92) | 1.40 (1.08, 1.95) | 0.240 |
| High-density lipoprotein cholesterol, mmol/L | 1.01 (0.87, 1.19) | 1.01 (0.90, 1.17) | 0.775 |
| Low-density lipoprotein cholesterol, mmol/L | 2.68 (2.09, 3.31) | 2.93 (2.32, 3.46) | 0.030 |
| Baseline GS | 20.0 (14.0, 28.0) | 25.0 (16.0, 31.5) | 0.007 |
| Follow-up GS | 21.0 (15.0, 28.0) | 26.5 (19.0, 31.5) | <0.001 |
| △GS | 0 (-1.0, 2.0) | 2.5 (1.0, 5.0) | <0.001 |
| Baseline 3V-QFR | 2.87 (2.80, 2.90) | 2.86 (2.79, 2.90) | 0.300 |
| Follow-up 3V-QFR | 2.87 (2.81, 2.91) | 2.76 (2.68, 2.83) | <0.001 |
| △3V-QFR | 0 (-0.02, 0.02) | -0.08 (-0.10, -0.06) | <0.001 |

Data are expressed as n (%), mean ± SD, or median (25th, 75th percentiles).

Abbreviations: PCI, percutaneous coronary intervention; ACEI, angiotensin-converting enzyme inhibitor; ARB, angiotensin II receptor antagonist; GS, Gensini score; QFR, quantitative flow ratio.

**Supplementary Figure 1.** **Study flow chart**

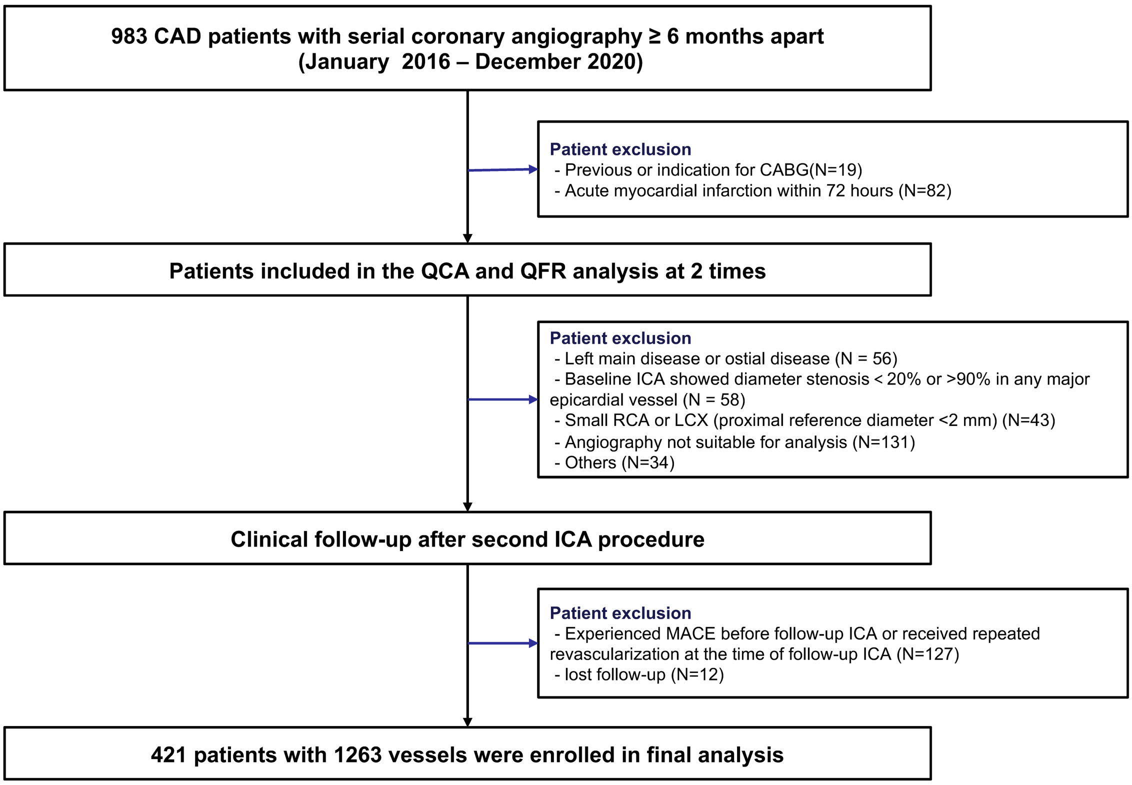


CAD, coronary artery disease; CABG, coronary artery bypass graft; QAG, quantitative coronary angiography; QFR, quantitative flow ratio; ICA, invasive coronary angiography; RCA, right coronary artery; LCX, left circumflex; MACE, major adverse cardiac events.

**Supplementary Figure 2. Receiver operator curves of 3V-QFR and ∆3V-QFR for discrimination of subsequent MACE.**

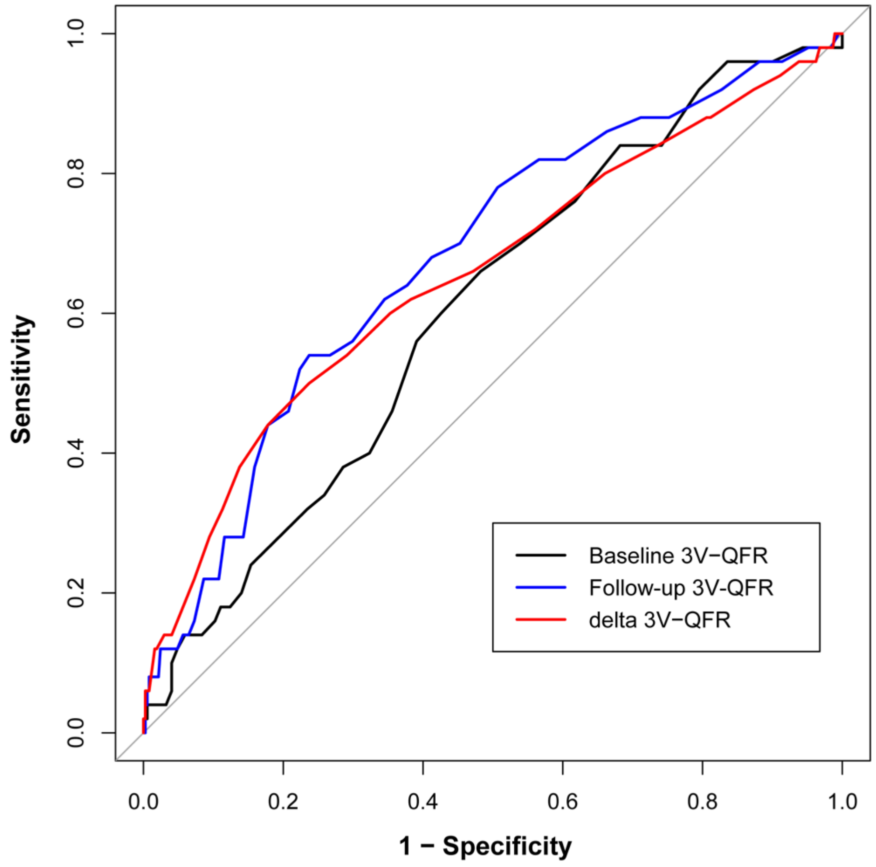


| Variable | AUC (95% CI) | Best cut-off | Sensitivity | Specitivity | Accuracy | p-value |
|---|---|---|---|---|---|---|
| **Baseline 3V-QFR** | 0.61 (0.53-0.69) | **2.86** | 0.66 | 0.52 | 0.53 | 0.008 |
| **Follow-up 3V-QFR** | 0.68 (0.60-0.76) | **2.78** | 0.54 | 0.76 | 0.74 | <0.001 |
| **Delta 3V-QFR** | 0.65 (0.57-0.74) | **-0.05** | 0.50 | 0.76 | 0.73 | <0.001 |

The black and blue lines are 3V-QFR based on baseline and follow-up angiograms, respectively; the red line is ∆3V-QFR. QFR, quantitative flow ratio; AUC, area under curve; CI: confidence interval; MACE, major adverse cardiac events.
